# Policy impact evaluation: A potential use case for longitudinal monitoring of viruses in wastewater at small geographic scales

**DOI:** 10.1101/2023.10.25.23297556

**Authors:** Elana M. G. Chan, Amanda Bidwell, Zongxi Li, Sebastien Tilmans, Alexandria B. Boehm

**Affiliations:** Department of Civil and Environmental Engineering, Stanford University, Stanford, California 94305, United States; Codiga Resource Recovery Center, Stanford University, Stanford, California 94305, United State

**Keywords:** Wastewater-based epidemiology, SARS-CoV-2, COVID-19, Quasi-experiment, Difference-in-differences, Policy evaluation, Causal inference

## Abstract

We used wastewater monitoring data to evaluate the impact of public health policies and interventions on the spread of COVID-19 among a university population. We first evaluated the correlation between incident, reported COVID-19 cases and wastewater SARS-CoV-2 RNA concentrations and observed changes to the correlation over time. Using a difference-in-differences approach, we evaluated the association between university COVID-19 policy changes and levels of SARS-CoV-2 RNA concentrations in wastewater. Policy changes associated with a significant change in campus wastewater SARS-CoV-2 RNA concentrations included changes to face covering recommendations, indoor gathering bans, and routine surveillance testing requirements and availability. We did not observe changes in SARS-CoV-2 RNA concentrations associated with other policy changes. The work presented herein demonstrates how longitudinal wastewater monitoring of viruses may be used for causal inference such as policy impact evaluation, especially at small geographic scales.

## 1.0 Introduction

Nonpharmaceutical interventions (NPIs) are actions that aim to reduce the spread of an infectious disease in a community—especially when the community has little immunity to the pathogen or a vaccine is not yet available.^1^ Examples of NPIs implemented widely in the United States at the start of the coronavirus disease 2019 (COVID-19) pandemic include face mask mandates, stay-at-home orders, non-essential business closures, and large gathering bans.^2^ Although the implementation of NPIs intended to benefit communities by flattening the epidemic curve—that is by reducing the peak number of cases and burden on the health care system— the implementation of NPIs also led to negative economic consequences and tolls on social well-being.^3–5^ Governments and institutional leadership are tasked with balancing public health, social well-being, and economic prospects in the face of epidemics. Evidence-based knowledge can help policymakers and leaders make better-informed decisions in dire situations.

Following the initial wave of the pandemic, several studies empirically assessed the impact of NPIs on health-related outcomes. The findings of these studies suggest that NPIs reduced the spread of severe acute respiratory disease syndrome coronavirus 2 (SARS-CoV-2) virus, with school and workplace closures, business restrictions, large gathering bans, and mask mandates among the most impactful NPIs.^6–10^ A review of the various methodologies used by these studies found that around half analyzed raw outcome data and half analyzed computed outcome data (i.e., raw outcome data was used to compute another outcome).^11^ The most common raw outcomes analyzed were clinical surveillance reports (e.g., confirmed cases or deaths) and human mobility (e.g., tracking of mobile phones).^11^ The most common computed outcomes analyzed were COVID-19 growth rate and effective reproduction number.^11^

Although clinical surveillance and mobile phone tracking are the most common sources of data used to evaluate NPIs, these data are not without biases and limitations. Counts of confirmed cases depend on clinical testing capacity and clinical testing rates, and deaths that occur outside of hospitals may be underreported.^6–8,10,11^ Furthermore, clinical testing behaviors have drastically changed with the availability of self-administered antigen tests which are not reported to health departments.^12^ Although mobility data through tracking of mobile phones is unaffected by changes in clinical testing, these data are biased towards individuals who opt into location history sharing and may not be a reliable proxy for SARS-CoV-2 transmission dynamics.^13,14^ Wastewater monitoring, which has gained heightened attention since the start of the COVID-19 pandemic, is a promising data source because it does not suffer some of the limitations of clinical surveillance and mobility data for epidemiological inference.

Wastewater monitoring refers to analyzing a sample of wastewater, which represents a pooled biological sample of the contributing population, for concentrations of infectious disease markers. Wastewater monitoring data capture contributions from both symptomatic and asymptomatic individuals and are not influenced by clinical testing availability or clinical test-seeking behaviors.^15^ Studies have reported that concentrations of SARS-CoV-2 RNA in wastewater solids are correlated with laboratory-confirmed incident COVID-19 cases—although this correlation has changed since the widespread availability of self-administered antigen tests.^16–19^ Several studies also demonstrated that wastewater monitoring can be used at geographic scales smaller than a sewershed (i.e., the population serviced by a wastewater treatment plant) to gain insight about COVID-19 incidence.^20–32^ A potential use case for wastewater monitoring at subsewershed scales is to assess public health policies.

The World Health Organization particularly suggests sampling at finer spatial scales when using wastewater monitoring data to inform targeted control interventions.^15^ Previous studies evaluating NPIs using clinical surveillance or mobility data were mostly conducted at national or subnational scales, and few of these studies investigated variation in the impact of NPIs on health-related outcomes among subpopulations.^11^ NPIs may be more or less impactful in a specific subpopulation compared to the general population (e.g., due to different interaction patterns) or public health goals may differ among subpopulations (e.g., universities aim to maximize on-campus activity).^33^ Wastewater monitoring data may be well-suited to objectively assess NPIs, particularly among subpopulations and despite low clinical testing rates.

In this study, we evaluate the potential use case of wastewater monitoring data to empirically assess the impact of NPIs on the spread of COVID-19 among a university population. We begin by assessing the correlation between wastewater concentrations of SARS-CoV-2 RNA and incident, reported COVID-19 cases at Stanford University and evaluate changes to this correlation over time. Next, we evaluate the association between policies implemented at Stanford University and changes in wastewater concentrations of SARS-CoV-2 RNA using a difference-in-differences (DiD) approach. DiD is a quasi-experimental design commonly used in econometrics—although it was first used in 1854 by John Snow for epidemiologic purposes— that aims to assess the impact of an intervention on an outcome but without the use of randomization.^34–36^ DiD designs have been used by several previous studies to evaluate the causal effects of COVID-19 policies on clinical or mobility outcomes.^37–44^

## 2.0 Methods

We used wastewater SARS-CoV-2 RNA monitoring data, COVID-19 case surveillance data, and dates associated with changes to campus COVID-19 policies. All calculations and statistical analyses were conducted in R (version 4.1.3). This study is approved by the Stanford Institutional Review Board (IRB) for human subject research (IRB-59746).

### 2.1 Wastewater monitoring data

We used wastewater monitoring data from the Codiga Resource Recovery Center (CR2C) and the Palo Alto Regional Water Quality Control Plant (RWQCP) for this analysis. CR2C is a pilot scale wastewater treatment facility that services a portion of the Stanford University campus (California, USA) including academic buildings and student and faculty housing (Figure S1 in the Supporting Information (SI)).^45,46^ CR2C services approximately 10,000 people with an estimated daily flow of approximately 0.5 million gallons of wastewater each day.^20,46^ CR2C is a subsewershed of the sewershed serviced by RWQCP which is operated by the City of Palo Alto (California, USA). RWQCP services approximately 236,000 people and treats approximately 20 million gallons of wastewater each day for Los Altos, Los Altos Hills, Mountain View, Palo Alto, Stanford University, and the East Palo Alto Sanitary District (Figure S1 in the SI).^47^

Prospective, longitudinal wastewater sampling from CR2C and RWQCP began July 2021 and October 2020, respectively, and is currently ongoing. Six samples per week are collected from CR2C; seven samples per week are collected from RWQCP. Sampling from CR2C was temporarily reduced to two samples per week between 1 November 2022 and 31 December 2022. Details about sampling and processing methods, including quality assurance and quality control metrics, are registered in protocols.io^48–50^ and have been described previously by Wolfe et al.^51^ and Boehm et al.^52^ A brief description is provided in the SI (Text S1).

For this analysis, we used concentrations of the SARS-CoV-2 RNA N gene in wastewater settled solids in gene copies (gc) per gram (g) dry weight (gc/g), both unnormalized (N) and normalized by pepper mild mottle virus (PMMoV) RNA concentrations in wastewater settled solids in gc/g (N/PMMoV). The N gene target is located near the frequently used N2 assay target,^53^ and we have confirmed no mutation in the target over the course of the pandemic.^19^

PMMoV is a commonly used marker of wastewater fecal strength, and based on a mass balance model N/PMMoV should scale with disease incidence rate.^18,54,55^ We used data between 29 July 2021 and 9 August 2023 (CR2C: 590 days; RWQCP: 736 days). The measured N gene concentration was below the limit of detection (approximately 1,000 gc/g) in 29 samples from CR2C. No samples from RWQCP were below the limit of detection. We imputed half the limit of detection (500 gc/g) for the N gene concentration for samples below the limit of detection. There were no non-detects for PMMoV in the dataset. Data from RWQCP between 16 November 2020 and 31 December 2022 have been published previously by Boehm et al.^52^ and are publicly available through the Stanford Digital Repository (https://doi.org/10.25740/cx529np1130).^56^ Data from CR2C have not yet been published. All wastewater monitoring data used in this study are publicly available through the Stanford Digital Repository (https://doi.org/10.25740/ch598gf0783).^57^

### 2.2 COVID-19 case surveillance data

Reported COVID-19 cases from student-reported, self-administered antigen tests and laboratory-based PCR tests among students residing in the CR2C subsewershed are available from Stanford University. We used case data between 29 July 2021 and 9 August 2023 for this analysis. Additional details of these daily case counts are provided in the SI (Text S2).

### 2.3 Campus COVID-19 policies

Dates and details of changes to Stanford University’s COVID-19 policies were obtained from Stanford COVID-19 Health Alerts.^58^ There were 15 unique dates on which campus COVID-19 policies changed (Table 1) during the study period. We categorized policies into three groups: masking (i.e., those involving the use of face coverings), mobility (i.e., those involving movement or gathering of individuals), and testing (i.e., those relating to laboratory-based surveillance testing). We further differentiated policies between those that enforced rules (i.e., restrictions) and those that relaxed existing rules (i.e., relaxations). More information about each policy is included in the SI (Table S1).

**Table 1.** Changes to COVID-19 policies at Stanford University since regular wastewater sampling began at the Codiga Resource Recovery Center.

| Date | Category | Type | Policy |
| --- | --- | --- | --- |
| 8/3/2021 | Masking | Restriction | Face coverings required indoors |
| 9/2/2021 | Masking;<br>Mobility | Restriction;<br>Restriction | Face coverings recommended outdoors in crowded settings; Indoor student parties prohibited |
| 9/20/2021 | Testing;<br>Mobility | Restriction;<br>Relaxation | Surveillance testing required for faculty, staff, and postdoctoral scholars regardless of vaccination status; Revised travel guidelines in effect |
| 10/8/2021 | Mobility | Relaxation | Indoor student parties can resume |
| 1/3/2022 | Mobility | Restriction | Online classes start |
| 1/5/2022 | Mobility;<br>Mobility | Restriction;<br>Restriction | No indoor events and gatherings; Outdoor-only gatherings and meetings prohibited |
| 1/18/2022 | Mobility | Relaxation | In-person instruction resumes |
| 1/21/2022 | Mobility | Relaxation | Outdoor-only gatherings and meetings allowed again |
| 1/28/2022 | Mobility | Relaxation | Indoor events and gatherings can resume |
| 3/2/2022 | Masking | Relaxation | Face coverings strongly recommended, regardless of vaccination status |
| 4/7/2022 | Testing | Relaxation | Surveillance testing no longer required for students |
| 10/24/2022 | Masking | Relaxation | Masks no longer required in classrooms and on campus shuttles |
| 1/6/2023 | Masking | Restriction | Face coverings strongly recommended indoors and in crowded outdoor settings |
| 3/24/2023 | Testing | Relaxation | Optional, free, laboratory-based PCR testing ends for employees |
| 6/18/2023 | Testing | Relaxation | Optional, free, laboratory-based PCR testing ends for students |

### 2.4 Correlation analysis

Incident COVID-19 cases within the CR2C subsewershed were reported daily while between 2– 6 wastewater samples per week were collected and analyzed from CR2C during the analysis period. Clinical case surveillance data may contain reporting biases on weekends. To compare the two time series, we calculated weekly average N concentrations, N/PMMoV concentrations, and incident COVID-19 cases for each epidemiological week (i.e., Sunday through Saturday). Neither raw nor log_10_-transformed weekly average N or N/PMMoV concentrations from CR2C are normally distributed (Shapiro-Wilk normality test, p < 0.01), so we used Kendall’s tau correlation to test the null hypothesis that weekly average wastewater SARS-CoV-2 RNA concentrations and weekly average incident COVID-19 cases in the CR2C subsewershed are not temporally correlated. We tested this null hypothesis using both unnormalized (N) and normalized (N/PMMoV) wastewater concentrations. We used the KendallTauB function from the DescTools R package to compute the 95% confidence interval for each tau estimate.^59^

We further conducted three subgroup correlation analyses. First, we grouped the data by whether wastewater sample or clinical specimen collection occurred during the academic year (autumn, winter, or spring quarter) or nonacademic year (summer quarter). We used the date halfway between the last day of classes of the previous quarter and first day of classes of the following quarter to define the start and end of quarters.^60^ Second, we grouped the data by whether wastewater sample or clinical specimen collection occurred before or after the requirement for laboratory-based surveillance testing was suspended for vaccinated and boosted students (7 April 2022) (Table 1). The laboratory-based surveillance testing program required fully vaccinated students to test once a week (twice a week for unvaccinated students) and therefore intended to capture both symptomatic and asymptomatic cases through routine testing. Third, we grouped the data by whether wastewater sample or clinical specimen collection occurred before or after 1 May 2022.^19^ This date represents a point in time when self-administered COVID-19 antigen tests, the results of which are not reportable to health departments, were widely available.^12,19^ For each subgroup, we grouped weekly average wastewater concentrations and incident case counts based on the end date of the epidemiological week. In total, we conducted 14 correlation analyses using subsets of the same sets of data to test the same null hypothesis, so we used an alpha value of 0.05 / 14 = 0.004 to account for multiple hypothesis testing when interpreting the p value associated with each tau estimate.

### 2.5 Policy impact evaluation

We used PMMoV-normalized wastewater concentrations for the remainder of the analysis as the correlation between incident COVID-19 cases and wastewater SARS-CoV-2 RNA concentrations were similar using N and N/PMMoV, and a mass balance model suggests the N/PMMoV ratio should scale with incidence rate.^55^ To assess the association between campus COVID-19 policies and changes in N/PMMoV measurements at CR2C, we used a difference-in-differences (DiD) approach. For the DiD design, we assumed that policies went into effect at midnight on the date of implementation (day = 0). We defined the pre-treatment period as the 14 days before a policy was implemented (days -14 to -1) and the post-treatment period as the 14 days after a policy was implemented (days 0 to 13). We chose 14 days because 14 days is the maximum incubation period for SARS-CoV-2 and people who shed SARS-CoV-2 RNA typically do so at the start of infection.^61–65^ We assumed that the RWQCP sewershed represents a reasonable comparison group for the CR2C subsewershed. With the exception of the East Palo Alto Sanitary District, RWQCP services cities in Santa Clara County which is the same county that Stanford University is located in. Santa Clara County entered the least restrictive “Yellow

Tier” of California’s Blueprint for a Safer Economy on 19 May 2021, which lifted most local orders.^66^ Moreover, California met the criteria under the Blueprint for a Safer Economy to fully reopen the economy on 15 June 2021.^67^ Regular sampling began at CR2C on 29 July 2021; therefore, we assumed that policies implemented by Stanford University thereafter (Table 1) were only applicable to the CR2C subsewershed population and not the greater RWQCP sewershed population. The two exceptions were 3 August 2021 and 2 March 2022 because Santa Clara County also issued the same policies (Table 1).^68,69^ Further justification for using RWQCP as a comparison group is included in the SI (Text S3).

We used a regression model to implement our DiD approach (Equation 1).^70^ A value of 0 for *time* represents the pre-treatment period (days -14 to -1) and a value of 1 represents the post-treatment period (days 0 to 13). A value of 0 for *treated* represents the untreated group (RWQCP) and a value of 1 represents the treated group (CR2C). The coefficient of the interaction between *time* and *treated* (*β*_3_) represents the DiD estimator, or the average treatment effect on the treated (ATT).^35,70^ In this study, a positive ATT value suggests that a policy was associated with an increase in wastewater N/PMMoV concentrations; a negative ATT value suggests that a policy was associated with a decrease in wastewater N/PMMoV concentrations.

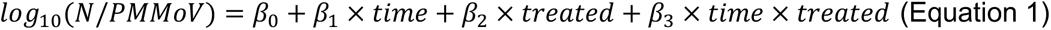

We recorded the ATT and associated p value for each policy in Table 1 except for the two policies that Santa Clara County also issued (see above). For each date corresponding to a policy change, we also recorded the trend in N/PMMoV over the past 14 days and the relative level of the 7-day right-aligned moving average of N/PMMoV on that date (see Text S4 in the SI for a description of trend and level calculations).^71^ R code for the DiD analysis is available through the Stanford Digital Repository (https://doi.org/10.25740/ch598gf0783).^57^

## 3.0 Results and Discussion

### 3.1 Correlation between wastewater concentrations of SARS-CoV-2 RNA and incident COVID-19 cases

Between 29 July 2021 and 9 August 2023, wastewater N gene concentrations from CR2C ranged from not detected to 2.4 × 10^6^ gc/g (mean: 1.3 × 10^5^ gc/g, median: 4.4 × 10^4^ gc/g) (Figure 1). PMMoV-normalized wastewater concentrations ranged from not detected to 5.0 × 10^-3^ (mean: 2.4 10^-4^, median: 6.4 × 10^-5^) (Figure 1). Reported daily incident COVID-19 cases within the CR2C subsewershed ranged from 0 cases to 420 cases (mean: 52 cases, median: 15 cases) (Figure 1). Over the entire analysis period (the week ending on 31 July 2021 through the week ending on 12 August 2023), weekly average wastewater SARS-CoV-2 RNA concentrations were positively and significantly correlated with weekly average incident COVID-19 cases using unnormalized N gene concentrations but not significantly when using normalized N gene concentrations (Table 2). The subgroup analyses suggest that the correlation between wastewater SARS-CoV-2 RNA concentrations and incident COVID-19 cases changed over time.

**Figure 1.**
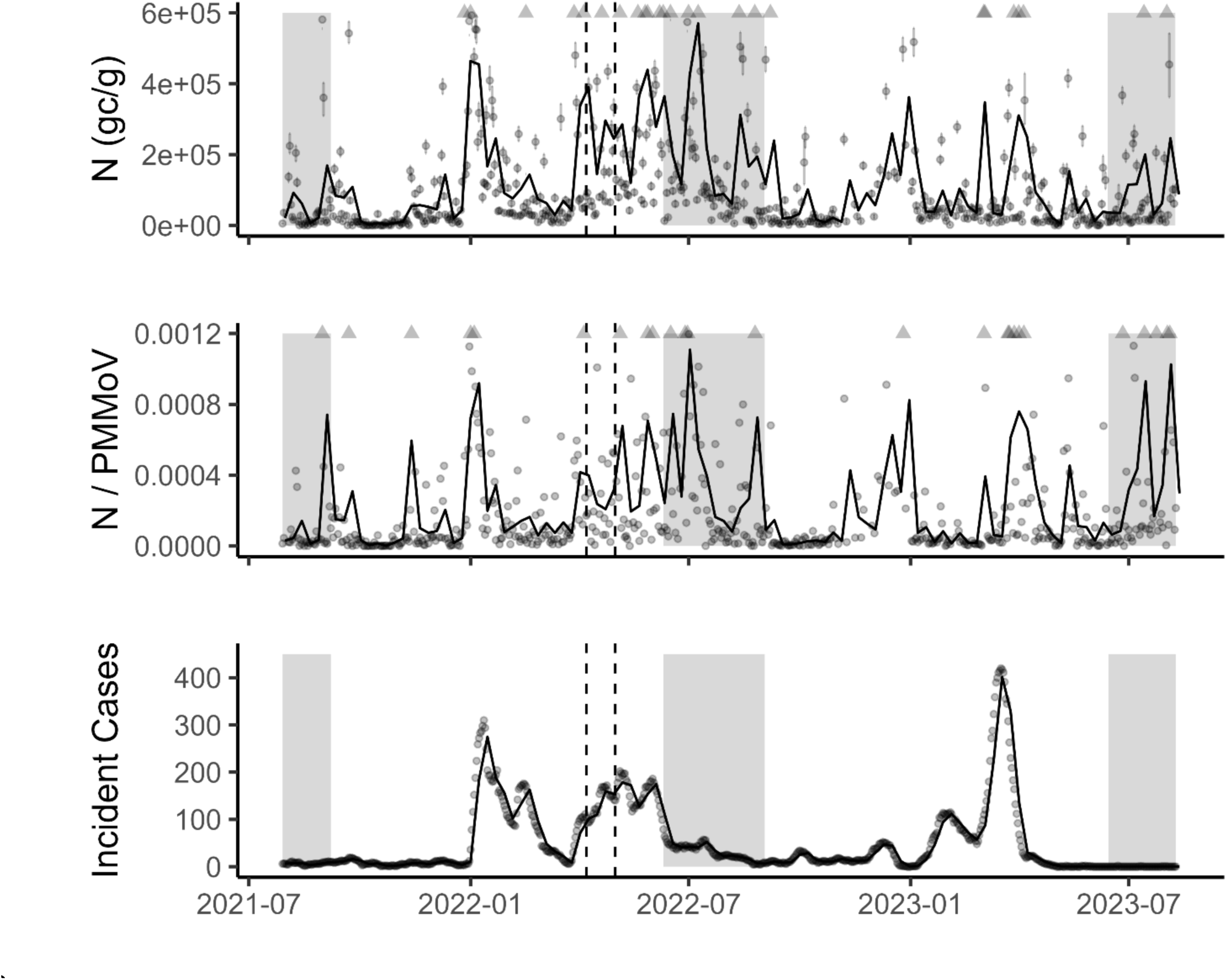
(Top) N gene concentrations in gene copies per dry gram dry weight (gc/g), (middle) N/PMMoV concentrations, and (bottom) incident COVID-19 cases over time in the Codiga Resource Recovery Center (CR2C) subsewershed. Gray circles represent measurements; error bars are one standard deviation. Gray triangles indicate measurements outside of the range shown on the plot. Black lines represent weekly average values. The shaded area corresponds to the nonacademic year. The dashed lines correspond to the date the surveillance testing requirement was suspended (7 April 2022) and the date of widespread availability of self-administered COVID-19 antigen tests in the region (1 May 2022).

Weekly average wastewater SARS-CoV-2 RNA concentrations were positively and significantly correlated with weekly average incident COVID-19 cases during the academic year using both unnormalized and normalized N gene concentrations; this correlation was insignificant during the nonacademic portion of the year (Table 2). The decrease in students on campus and increase in nonresidential visitors during the nonacademic portion of the year may explain the insignificant correlation during the nonacademic year. The COVID-19 case surveillance data only includes reported student cases residing within the CR2C subsewershed, but infected, nonresidential visitors may still contribute viral RNA to the wastewater that flows to CR2C.

**Table 2.**
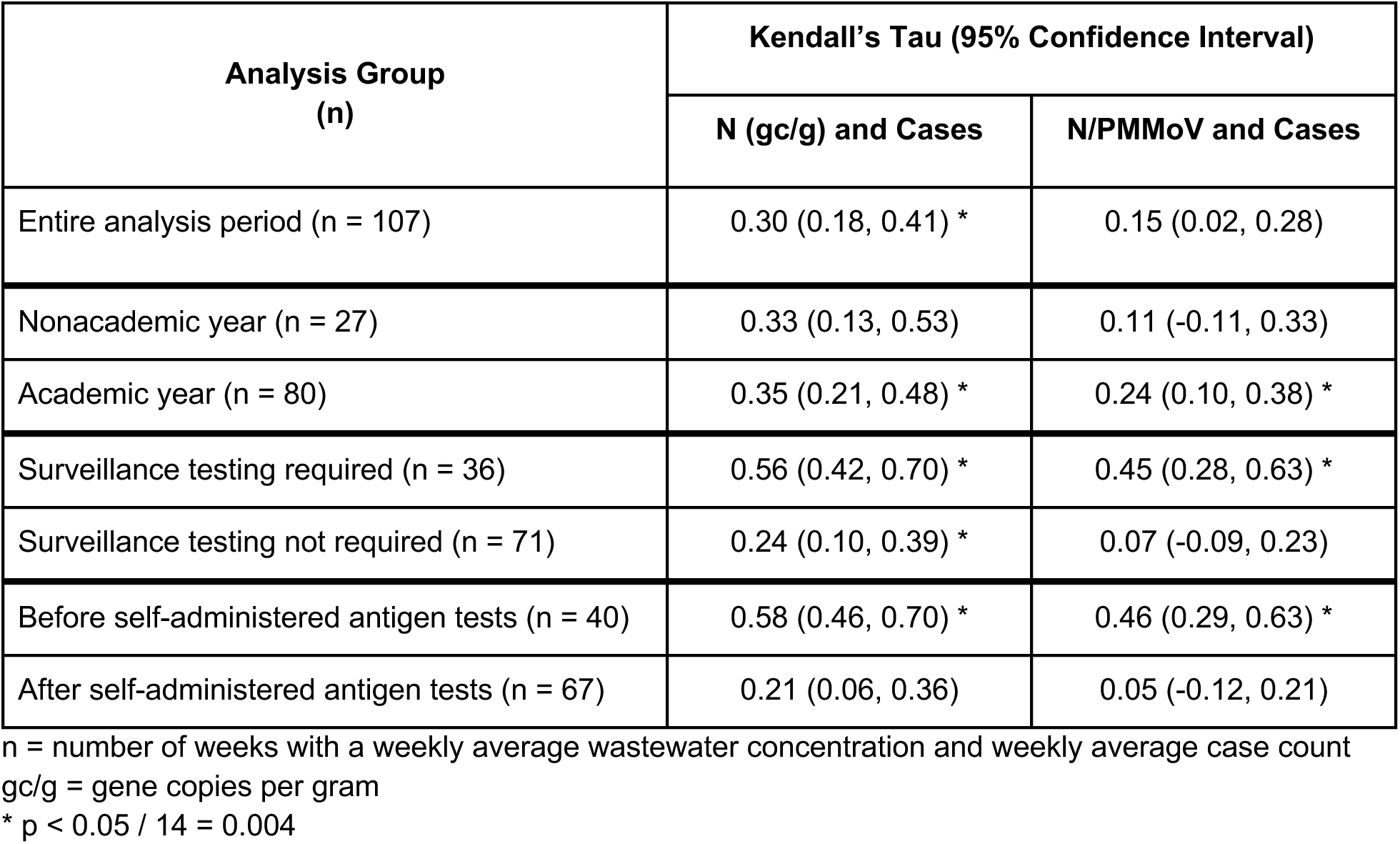
Kendall’s tau correlation between weekly average wastewater SARS-CoV-2 RNA concentrations and incident COVID-19 cases within the Codiga Resource Recovery Center subsewershed

Weekly average wastewater SARS-CoV-2 RNA concentrations were positively and significantly correlated with weekly average incident COVID-19 cases before the suspension of surveillance testing using both unnormalized and normalized N gene concentrations; this correlation was insignificant after the suspension of surveillance testing using normalized N gene concentrations only (Table 2). The required, laboratory-based surveillance testing program intended to capture both symptomatic and asymptomatic cases through routine testing. Thus, fewer asymptomatic cases may have been captured in the case data after surveillance testing was suspended which may explain the insignificant correlation after this policy change.

Lastly, weekly average wastewater SARS-CoV-2 RNA concentrations were positively and significantly correlated with weekly average incident COVID-19 cases before the widespread availability of self-administered antigen tests using both unnormalized and normalized N gene concentrations; this correlation was insignificant after the widespread availability of self-administered antigen tests (Table 2). Positive, laboratory-based PCR tests are reportable under state-disease reporting laws;^72^ however, self-reporting of self-administered antigen test results is voluntary. The widespread availability of self-administered antigen tests may have contributed to underreporting of cases which may explain the insignificant correlation after the change in testing options.

It is not possible to deduce the main driver for the change in correlation between wastewater SARS-CoV-2 RNA concentrations and incident COVID-19 cases over time, but we suspect that the change is due to several factors including changes in routine COVID-19 surveillance testing requirements, changes in test reporting, and overall decreases in test-seeking behaviors as the pandemic continues.^19,73,74^ Wastewater monitoring data are independent of test-seeking behaviors or test reporting patterns so may be a less biased tool for monitoring public health, particularly in periods characterized by low test-seeking and reporting rates.

### 3.2 Association between campus COVID-19 policies and changes in wastewater concentrations of SARS-CoV-2 RNA

Because the reliability of campus COVID-19 case surveillance data has changed over the course of the pandemic, we used wastewater monitoring data from CR2C to evaluate the impact of COVID-19 policies at Stanford University using a DiD approach. Table 3 summarizes the average treatment effect on the treated (ATT) and associated p value for each unique date associated with a change in campus COVID-19 policies as estimated using Equation 1. The two policies that were also implemented by Santa Clara County were omitted from the analysis.

**Table 3.** Difference-in-differences analysis to evaluate the association between campus COVID-19 policies and changes in wastewater N/PMMoV concentrations.

| Date<br>(Trend/Level) | Policy | ATT<br>(p value) |
| --- | --- | --- |
| 9/2/2021<br>(Upward/High) | Face coverings recommended outdoors in crowded settings<br>Indoor student parties prohibited | 0.72<br>(0.02) |
| 9/20/2021<br>(None/Medium) | Surveillance testing required for faculty, staff, and postdoctoral scholars regardless of vaccination status<br>Revised travel guidelines in effect | -0.61<br>(0.05) |
| 10/8/2021<br>(None/Low) | Indoor student parties can resume | -0.42<br>(0.2) |
| 1/3/2022<br>(Upward/High) | Online classes start | -0.078<br>(0.8) |
| 1/5/2022<br>(Upward/High) | No indoor events and gatherings<br>Outdoor-only gatherings and meetings prohibited | -0.16<br>(0.6) |
| 1/18/2022<br>(None/High) | In-person instruction resumes | -0.00048<br>(1.0) |
| 1/21/2022<br>(None/High) | Outdoor-only gatherings and meetings allowed again | -0.11<br>(0.6) |
| 1/28/2022<br>(Downward/Low) | Indoor events and gatherings can resume | 0.47<br>(0.03) |
| 4/7/2022<br>(None/High) | Surveillance testing no longer required for students | -0.62<br>(0.02) |
| 10/24/2022<br>(None/Low) | Masks no longer required in classrooms and on campus shuttles | 0.11<br>(0.7) |
| 1/6/2023<br>(Downward/Medium) | Face coverings strongly recommended indoors and in crowded outdoor settings | -0.26<br>(0.3) |
| 3/24/2023<br>(Upward/High) | Optional, free, laboratory-based PCR testing ends for employees | 0.72<br>(0.008) |
| 6/18/2023<br>(None/Low) | Optional, free, laboratory-based PCR testing ends for students | 0.37<br>(0.3) |

ATT = average treatment effect on the treated

Dates associated with a significant changes (p ≤ 0.05) in wastewater N/PMMoV concentrations are shaded (red if ATT > 0 and blue if ATT < 0). For all dates, the 14-day trend in N/PMMoV and the 14-day trend in N/PMMoV and the relative level of the 7-day right-aligned moving average of N/PMMoV is provided. In total, we analyzed thirteen unique dates on which at least one change in campus COVID-19 policies went into effect. Most policy change dates were not associated with a significant change in wastewater N/PMMoV concentrations at CR2C. We expected policy relaxations to be associated with no significant change in wastewater N/PMMoV concentrations because these policy types are not intended to curb virus transmission. Meanwhile, we expected policy restrictions to be associated with a significant decrease in wastewater N/PMMoV concentrations because these policy types are intended to curb virus transmission. Five policy change dates were associated with a significant change in wastewater N/PMMoV concentrations (Table 3; Figure S2 in the SI). Three of these dates corresponded to policy relaxations, one corresponded to a policy restriction, and one corresponded to both a policy relaxation and restriction, and these policies included all categories (masking, mobility, testing). A depiction of the DiD approach using the date when indoor events and gatherings were allowed to resume (28 January 2022) as an example is shown in the SI (Figure S3).

The policy restriction associated with a significant change in N/PMMoV concentrations (recommending face coverings outdoors and prohibiting indoor parties on 2 September 2021) was associated with a significant increase rather than decrease in N/PMMoV concentrations. N/PMMoV concentrations were already exhibiting a significant upward trend on 2 September 2021. These policy restrictions may have been triggered by a rise in infections on campus, so N/PMMoV concentrations may have continued to increase over the following 14 days as part of the natural epidemic curve.^75^ Similarly, the significant increase in N/PMMoV concentrations associated with the ending of optional, free, laboratory-based PCR testing for employees (24 March 2023) may be because N/PMMoV concentrations were already exhibiting a significant upward trend on 24 March 2023. N/PMMoV concentrations may have continued to increase over the following 14 days as part of the natural epidemic curve and not because of this policy relaxation.^75^ The significant increase in N/PMMoV concentrations associated with allowing indoor gatherings to resume (28 January 2022) may suggest that indoor gatherings are high-risk activities for SARS-CoV-2 transmission on campus. The significant decrease in N/PMMoV concentrations associated with suspending the surveillance testing requirement for students (7 April 2022) is difficult to reconcile with expectations. Given our expectation about policy relaxations, we did not expect this policy relaxation to be associated with a significant change in N/PMMoV concentrations but there are several other factors that can affect the DiD results as discussed later with limitations of the DiD design. The remaining date (20 September 2021) was associated with a significant decrease in N/PMMoV concentrations. Both a policy relaxation (revised travel guidelines) and policy restriction (surveillance testing required for all faculty, staff, and postdoctoral scholars) were implemented; it is not possible to disentangle the individual causal effects of policies implemented on the same day.

Because the wastewater data from CR2C and RWQCP are not truly independent—although CR2C comprises only a very small proportion of RWQCP—we also implemented the DiD analysis using wastewater data from the San José-Santa Clara Regional Wastewater Facility (RWF)^76^ as a comparison group (Table S2 in the SI) and found similar results as provided above (see SI). The similar findings using a different comparison group strengthens the credibility of our DiD analysis and affirms the plausibility of the parallel trends assumption.^77^

There are several limitations to our DiD analysis which may impact the interpretation of our results. First, policies may not be associated with immediate effects on outcomes.^75^ The policies we considered may have been associated with long-term effects on wastewater N/PMMoV concentrations despite being associated with null short-term effects. However, we determined that the 14 days preceding and succeeding a policy was the most justified time interval for the DiD design given that the maximum incubation period for SARS-CoV-2 is 14 days and people who shed SARS-CoV-2 RNA generally do so at the start of infection.^61–65^ When using 14 days, the pre- or post-treatment period of one policy overlapped with part of the pre- or post-treatment period of another policy for policies implemented close together which may lead to cumulative impacts on wastewater N/PMMoV concentrations that are not possible to disentangle. Second, COVID-19 vaccines—which are a highly effective pharmaceutical intervention—were widely available during our entire analysis period with vaccination rates being very high among the Stanford University and greater Santa Clara County populations.^68,78^ The insignificant change in N/PMMoV concentrations we observed after most policy restrictions may be due to the effectiveness of vaccines and not necessarily because these policies are unimportant. Modeling studies suggest that the success of COVID-19 vaccination programs is contingent upon appropriate use of NPIs.^79–82^ Third, there were sometimes campus announcements or national news headlines about COVID-19 preceding the implementation of policies which could impact peoples’ behaviors leading up to the actual policy change date.^75^ Peoples’ knowledge about the gravity of the COVID-19 pandemic has been shown to influence the effectiveness of lockdown policies.^39^ Lastly, the large influx of students and visitors to campus at the starts of quarters and commencements may mask the intended effects of campus policies implemented around the same time. Unlike major holidays, which result in nationwide travel (i.e., to both CR2C and RWQCP), commencements and the first day of classes result in migration specifically to Stanford’s campus that is unaccounted for in the DiD design. We conducted the DiD analysis for commencements and the first day of classes of each quarter (Table S2 in the SI) and only two starts of quarters were associated with a significant change in wastewater N/PMMoV concentrations at CR2C (decrease at the start of autumn 2021 and increase at the start of spring 2022).

Nonetheless, our analysis is highlighted by several strengths. We empirically assessed the impact of policies among a vaccinated university population. A study focused on vaccinated populations at universities in the United Kingdom concluded that mask wearing and social distancing measures were most important using a modeling approach.^33^ Our approach using empirical data suggests that most policies were associated with insignificant impacts although gatherings may be a particularly high-risk factor for virus transmission on campus. For our empirical approach, we used longitudinal wastewater monitoring data. We first demonstrated that the correlation between wastewater SARS-CoV-2 RNA concentrations and incident COVID-19 cases at Stanford has changed over time, suggesting that wastewater monitoring data is more reliable than clinical surveillance data for use cases such as policy impact evaluation. We further considered both policy restrictions and policy relaxations during a period when COVID-19 vaccines were widely available. Previous studies that empirically assessed the impact of NPIs on health-related outcomes generally only focused on restrictions and were most commonly conducted at the start of the pandemic when economies were not fully opened and vaccines were not available. It is not only important to evaluate the implementation of policies but also whether policies are eventually relaxed appropriately, especially because the success of vaccination campaigns is influenced by NPIs.^79–82^ Lastly, we used a quasi-experimental design to statistically evaluate changes in wastewater N/PMMoV concentrations associated with policy change dates rather than simply using a qualitative or before-and-after descriptive approach. The DiD design presented herein may be adapted in the future to assess the implications of NPIs for other diseases that may be monitored in wastewater such as influenza and respiratory syncytial virus.^83^

## 4.0 Conclusions

We assessed the correlation between wastewater concentrations of SARS-CoV-2 RNA and incident, reported COVID-19 cases at a university and evaluated changes to this correlation over time. Consistent with other studies, we provide evidence that the correlation between wastewater SARS-CoV-2 RNA concentrations and incident COVID-19 cases has changed over time. We further investigated the use of longitudinal wastewater monitoring data for policy impact evaluation. Using a DiD approach, we observed that most dates on which changes to campus COVID-19 policies occurred were not associated with a significant change in wastewater SARS-CoV-2 RNA concentrations on campus. However, we did not expect to observe a significant change in wastewater SARS-CoV-2 RNA concentrations after all policy change dates, particularly those dates on which existing policies were phased out. The work presented herein demonstrates how longitudinal wastewater monitoring of viruses may be used for causal inference such as policy impact evaluation, especially at small geographic scales.

## Data Availability

All data produced are available online at https://doi.org/10.25740/ch598gf0783.

## Supporting Information

Supporting Information: Supplementary figures, text, and tables, including sewershed maps and experimental details (PDF)

## Acknowledgements

We acknowledge feedback on study design, implementation, and interpretation from James Jacobs, Julie Parsonnet, Russell Furr, Rich Wittman, Robyn Tepper, Jorge Salinas, Bonnie Maldonado, Christina Kong, and Stephanie Kalfayan. We acknowledge Palo Alto and San Jose wastewater treatment plant staff and the CR2C Student Operators team for wastewater sample collection.

## For Table of Contents Only

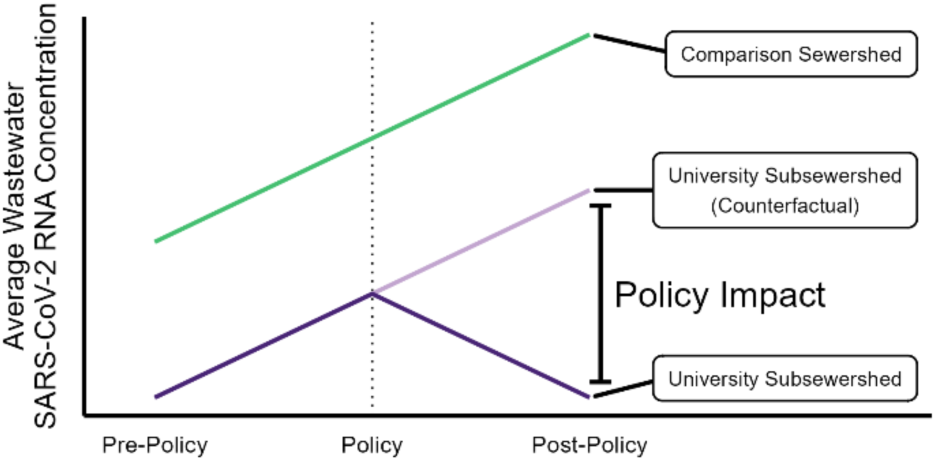

