## Supporting Information for "Policy impact evaluation: A potential use case for longitudinal monitoring of viruses in wastewater at small geographic scales"

**Figure S1. Map of wastewater treatment plant service areas** **2**

**Text S1. Wastewater solids sampling and processing methods** **3**

**Table S1. Detailed information about campus COVID-19 policies** **4**

**Text S2. Further information about COVID-19 surveillance data** **9**

**Text S3. Further justification for the difference-in-differences design** **10**

**Text S4. Description of wastewater trend and relative level calculations** **11**

**Figure S2. Wastewater N/PMMoV measurements at CR2C and RWQCP with difference-in-differences results** **12**

**Figure S3. Depiction of the difference-in-differences approach** **13**

**Table S2. Difference-in-difference analysis for campus COVID-19 policies and events using two different comparison groups** **14**

**References** **16**

**Figure S1. Map of wastewater treatment plant service areas**

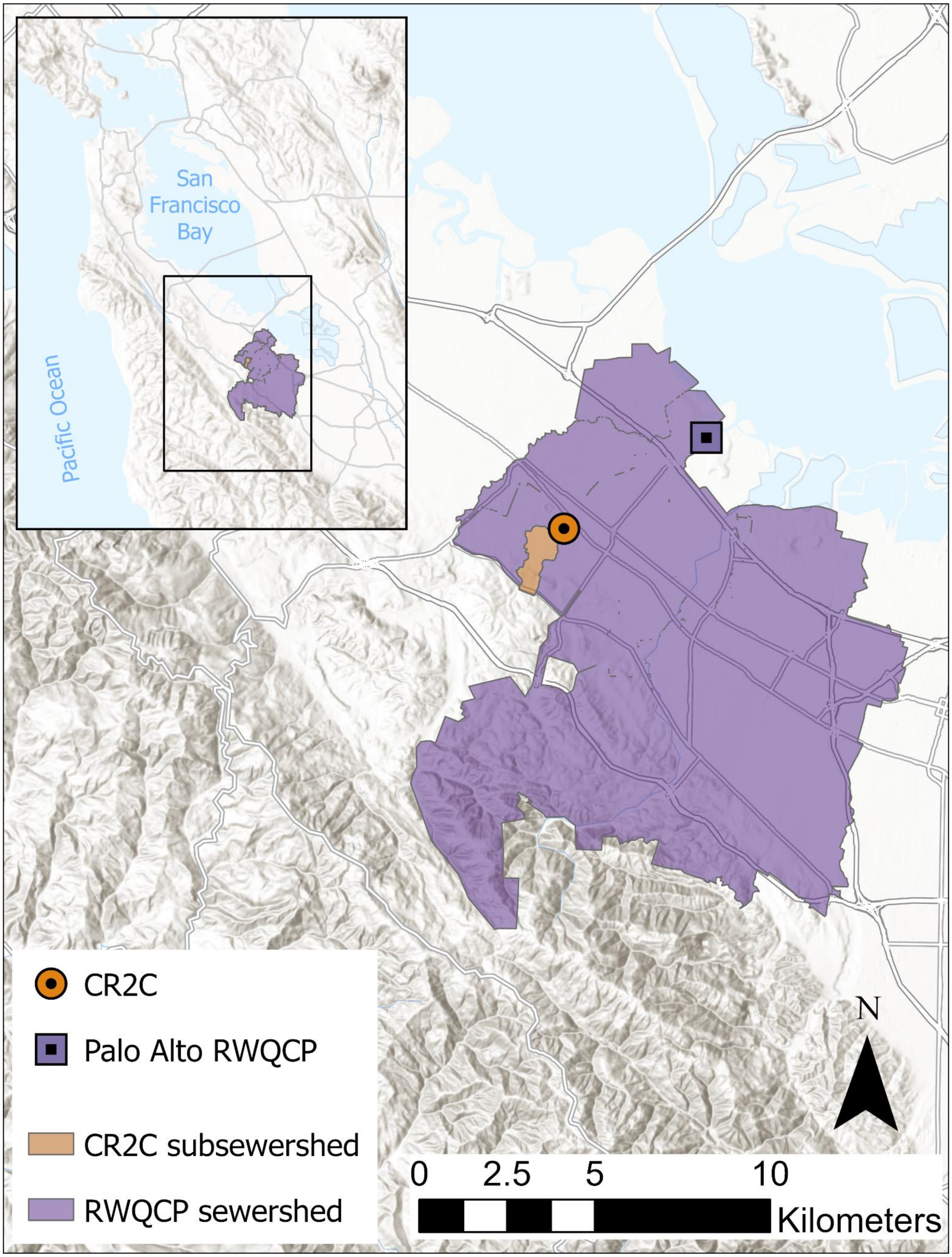


Figure S1. Service area of the Codiga Resource Recovery Center (CR2C) and Palo Alto Regional Water Quality Control Plant (RWQCP). Map created in ArcGIS Pro 3.1.1 using "Terrain with Labels" basemap.[1]

**Text S1. Wastewater solids sampling and processing methods**

Wastewater settled solids are collected from both the Codiga Resource Recovery Center (CR2C) and the Palo Alto Regional Water Quality Control Plant (RWQCP) for laboratory processing. Settled solids samples at CR2C are generated from a 24-hour composite sample of the wastewater influent that is allowed to settle. Settled solids samples at RWQCP are “grab” samples from the primary clarifier; these samples are essentially composite samples because solids in the primary clarifier collect over 12–24 hours.[2] The pre-analytical and analytical methods used to measure the RNA targets are registered[3–5] and have been previously outlined in peer-reviewed publications,[6,7] so they are not repeated herein. Measurements and reporting in those other publications follow Environmental Microbiology Minimal Information (EMMI) guidelines.

**Table S1. Detailed information about campus COVID-19 policies**

| Date | Category | Type | Policy | Description | Source |
|---|---|---|---|---|---|
| 8/3/2021 | Masking | Restriction | Face coverings required indoors | Santa Clara and San Mateo counties joined others in the Bay Area in issuing a requirement for face coverings indoors – regardless of an individual's COVID-19 vaccination status – effective tonight at midnight. In compliance with this new rule, all Stanford employees, students and visitors will be required to wear face coverings in indoor spaces. Exceptions are allowed for individuals who are alone in a private office or room or actively eating or drinking. | https://healthalerts.stanford.edu/covid-19/2021/08/02/face-coverings-for-indoor-campus-spaces/ |
| 9/2/2021 | Masking | Restriction | Face coverings recommended outdoors in crowded settings | In addition to the indoor mask requirement already in place, we are strongly recommending masking outdoors in crowded settings when 6 feet of distance from others cannot consistently be maintained. | https://healthalerts.stanford.edu/covid-19/2021/09/02/covid-19-update-and-new-protocols/ |
| | Mobility | Restriction | Indoor student parties prohibited | Indoor student parties will be prohibited until October 8, the end of the third week of the fall quarter for most students, to help limit the potential for virus transmission in this period when we are returning on-site. | |

| | | | | | |
|---|---|---|---|---|---|
| 9/20/2021 | Testing | Restriction | Surveillance testing required, regardless of vaccination status | Fully vaccinated faculty, staff and postdoctoral scholars will be required to test once a week. This is already the policy for fully vaccinated students but is a new requirement for these other groups. Twice weekly testing will be required for unvaccinated faculty, staff, postdoctoral scholars and students (who are currently required to test once a week). Testing will be once weekly for those who come to work on-site three days or fewer per week and are not fully vaccinated. | https://healthalerts.stanford.edu/covid-19/2021/08/27/update-on-covid-testing-travel-and-employee-support/ |
| | Mobility | Relaxation | Revised travel guidelines in effect | Fully vaccinated faculty, staff, postdocs and graduate students will be permitted to travel domestically and internationally without requesting a travel exception. Unvaccinated faculty, staff, postdocs and graduate students will be permitted to travel domestically but must be approved for a travel exception prior to commencing international travel. Travel associated with the Bing Overseas Studies Program, Stanford in New York and Stanford in Washington will be permitted for fully vaccinated undergraduates. In addition, undergraduates may travel within California for Stanford-related course field research and field trips that are accompanied by an instructor. The prohibition on other international travel and domestic travel outside California will remain in place. | |
| 10/8/2021 | Mobility | Relaxation | Indoor student parties can resume | Indoor student parties will be prohibited until October 8, the end of the third week of the fall quarter for most students, to help limit the | https://healthalerts.stanford.edu/covid-19/2021/09/02/covid-19- |

| | | | | | |
|---|---|---|---|---|---|
| | | | | potential for virus transmission in this period when we are returning on-site. | update-and-new-protocols/ |
| 1/3/2022 | Mobility | Restriction | Online classes start | Classes at Stanford will be online for the first two weeks of the winter quarter. The quarter will still begin Jan. 3 for most students, as scheduled. We will resume in-person instruction Tuesday, Jan. 18. | https://news.stanford.edu/report/2021/12/16/first-two-weeks-winter-classes/ |
| 1/5/2022 | Mobility | Restriction | No indoor events and gatherings | Essential academic and administrative meetings and trainings can continue, but conferences, social events, religious services and other gatherings generally should be moved outdoors, moved online or rescheduled. Outdoor events will require masking and should provide for social distancing. | https://healthalerts.stanford.edu/covid-19/2022/01/05/short-term-updates-to-campus-policies/ |
| | Mobility | Restriction | Outdoor-only gatherings and meetings prohibited | For students, outdoor-only gatherings and meetings of student organizations will be allowed beginning Jan. 21, as previously announced. Indoor house meetings and private residential gatherings continue to be allowed, with face coverings. | |
| 1/18/2022 | Mobility | Relaxation | In-person instruction resumes | Classes at Stanford will be online for the first two weeks of the winter quarter. The quarter will still begin Jan. 3 for most students, as scheduled. We will resume in-person instruction Tuesday, Jan. 18. | https://news.stanford.edu/report/2021/12/16/first-two-weeks-winter-classes/ |
| 1/21/2022 | Mobility | Relaxation | Outdoor-only gatherings and meetings allowed again | For students, outdoor-only gatherings and meetings of student organizations will be allowed beginning Jan. 21, as previously | https://healthalerts.stanford.edu/covid-19/2022/01/05/short-term- |

| | | | | | |
|---|---|---|---|---|---|
| | | | | announced. Indoor house meetings and private residential gatherings continue to be allowed, with face coverings. | updates-to-campus-policies/ |
| 1/28/2022 | Mobility | Relaxation | Indoor events and gatherings can resume | From now until Friday, Jan. 28, essential academic and administrative meetings and trainings can continue, but conferences, social events, religious services and other gatherings generally should be moved outdoors, moved online or rescheduled. Outdoor events will require masking and should provide for social distancing. | https://healthalerts.stanford.edu/covid-19/2022/01/05/short-term-updates-to-campus-policies/ |
| 3/2/2022 | Masking | Relaxation | Face coverings strongly recommended, regardless of vaccination status | With certain exceptions, face coverings will no longer be required but will continue to be strongly recommended on-site, regardless of vaccination status. In classrooms, face coverings will continue to be required through the beginning of spring quarter. However, individuals may remove face coverings while speaking. And, in compliance with State of California protocols, masking will still be required in these settings, regardless of vaccination status: Public transportation, including Marguerite buses. Healthcare facilities, including Vaden Health Center and Stanford hospitals and clinics. Childcare facilities (through March 11, after which masks remain strongly recommended). | https://healthalerts.stanford.edu/covid-19/2022/03/01/revised-masking-guidelines/ |
| 4/7/2022 | Testing | Relaxation | Surveillance testing no longer required | The requirement for COVID surveillance testing of vaccinated and boosted students is being suspended. | https://healthalerts.stanford.edu/covid-19/2022/04/07/guidance-on-covid-19-masking-and-testing/ |

| | | | | | |
|---|---|---|---|---|---|
| 10/24/2022 | Masking | Relaxation | Masks no longer required in classrooms and on Marguerite shuttles | With the level of COVID-19 cases on campus and in the surrounding region remaining low, Stanford's requirements for masking in classrooms and on Marguerite shuttles will be eased effective next Monday, Oct. 24. | https://healthalerts.stanford.edu/covid-19/2022/10/17/masking-requirements-boosters/ |
| 1/6/2023 | Masking | Restriction | Face coverings strongly recommended indoors and in crowded outdoor settings | Stanford strongly recommends wearing masks in classrooms and instructional spaces during the remainder of January. Stanford also strongly recommends masking indoors and in crowded outdoor settings. Masking is still required in healthcare facilities. | https://healthalerts.stanford.edu/covid-19/2023/01/06/classroom-masking-recommendations/ |
| 3/24/2023 | Testing | Relaxation | Free, laboratory-based PCR testing ends for employees | Free rapid tests will continue to be provided | https://healthalerts.stanford.edu/covid-19/2023/03/08/changes-to-covid-testing-program/ |
| 6/18/2023 | Testing | Relaxation | Free, laboratory-based PCR testing ends for students | Free rapid tests will continue to be provided | https://healthalerts.stanford.edu/covid-19/2023/03/08/changes-to-covid-testing-program/ |

**Text S2. Further information about COVID-19 surveillance data**

Daily COVID-19 case counts correspond to the date of positive test specimen collection. Both student-reported self-administered antigen tests and laboratory-based PCR tests through the university's surveillance testing program are included in the campus case surveillance data. As part of the university's surveillance testing program, vaccinated students were required to test once per week (twice per week for unvaccinated students) through 7 April 2022. Free, optional laboratory-based PCR testing continued to be available for students through 18 June 2023, so any cases thereafter were exclusively from student-reported self-obtained tests. The Codiga Resource Recovery Center (CR2C) subsewershed also includes faculty and staff housing, but nonstudents residing in the CR2C subsewershed are not included in the university's case data. Data provided by the state of California through data use agreements did not identify any COVID-19 cases in nonstudent housing areas during our entire analysis period.

**Text S3. Further justification for the difference-in-differences design**

A challenge associated with empirically evaluating policies and interventions—whether using clinical surveillance, mobility, or wastewater monitoring data—is that conducting a randomized controlled trial is generally not possible or ethical. Quasi-experimental designs, which aim to assess the impact of an intervention on an outcome but without the use of randomization, are therefore central in health policy impact evaluation.[8] Difference-in-differences (DiD) is one quasi-experimental design that is commonly used in econometrics—although it was first used in 1854 by John Snow for epidemiologic purposes.[9] The DiD design compares changes in an outcome before versus after a treatment between a treated and untreated group (treatment is not randomly assigned). The idea of the DiD design is that the treated and untreated groups follow the same time trends in the absence of treatment. Thus, the counterfactual outcome for the treated group after the treatment may be approximated to estimate the effect of the treatment on the treated group.[9,10]

In this study, we assumed the Palo Alto Regional Water Quality Control Plant (RWQCP) sewershed represents a reasonable "untreated" comparison group for the Codiga Resource Recovery Center (CR2C) subsewershed for reasons mentioned in the main text. Prior to implementing the DiD analysis, we compared raw wastewater N/PMMoV concentrations between the CR2C subsewershed and RWQCP sewershed on days that did not fall within a post-treatment period of a policy to further evaluate the use of RWQCP as a comparison group. We used Kendall's tau correlation test as neither raw nor log10-transformed N/PMMoV values from CR2C and RWQCP are normally distributed (Shapiro-Wilk normality test, $p < 0.01$). $Log_{10}$-transformed wastewater N/PMMoV concentrations were positively correlated over time between CR2C and RWQCP outside of post-treatment periods (tau = 0.19, $p < 0.001$, N = 384 days); we assume this demonstrates that CR2C and RWQCP exhibit the same time trends in the absence of policies and satisfies the parallel trends assumption of the DiD design.[9]

**Text S4. Description of wastewater trend and relative level calculations**

For each date corresponding to a policy change, we recorded the trend in N/PMMoV over the past 14 days and the relative level of the 7-day right-aligned moving average of N/PMMoV on the date alongside the results from the difference-in-differences (DiD) analysis. Trends were calculated using the percent change method described by Chan et al.[11] “Upward” indicates any significant upward trend, “None” indicates no significant trend, and “Downward” indicates any significant downward trend in N/PMMoV over the past 14 days on the date. Relative levels were determined retrospectively from all 7-day right-aligned moving average N/PMMoV values from CR2C in our analysis period. “Low”, “Medium”, and “High” indicate that the 7-day right-aligned moving average of N/PMMoV on the date was in the bottom, middle, and top tertile, respectively. Note that $log_{10}$-transformed raw values of N/PMMoV were used for the DiD analysis (Equation 1 in the main text); moving average values of N/PMMoV were only used to classify wastewater N/PMMoV concentrations as relatively high, medium, or low on a given day.

**Figure S2. Wastewater N/PMMoV measurements at CR2C and RWQCP with difference-in-differences results**

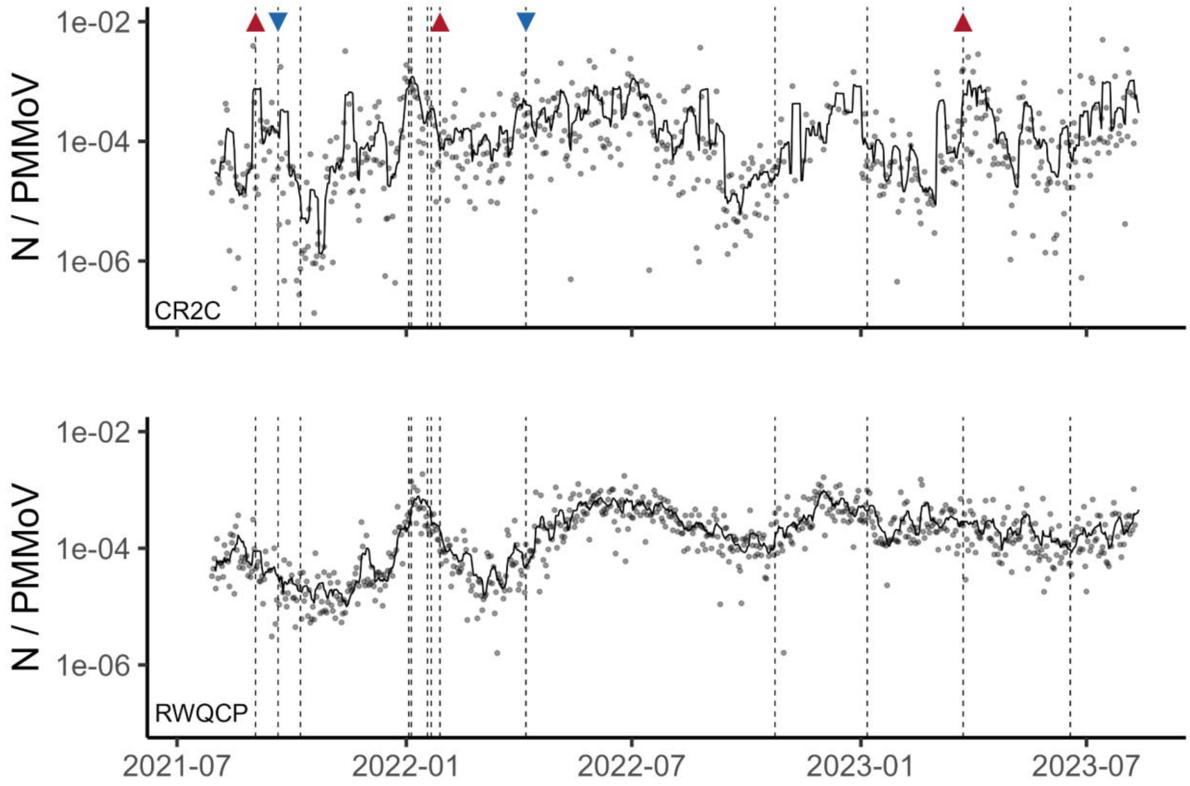


Figure S3. Wastewater N/PMMoV concentrations over the entire analysis period from (top) the Codiga Resource Recovery Center (CR2C) and (bottom) the Palo Alto Regional Water Quality Control Plant (RWQCP). Concentrations are displayed on a $\log_{10}$ scale with a 7-day right-aligned moving average line. Dashed lines correspond to the dates of COVID-19 policy changes at Stanford University. Triangles denote policy change dates associated with a significant change in N/PMMoV concentrations at CR2C based on the difference-in-difference analysis (red, up-pointing = increase; blue down-pointing = decrease).

**Figure S3. Depiction of the difference-in-differences approach**

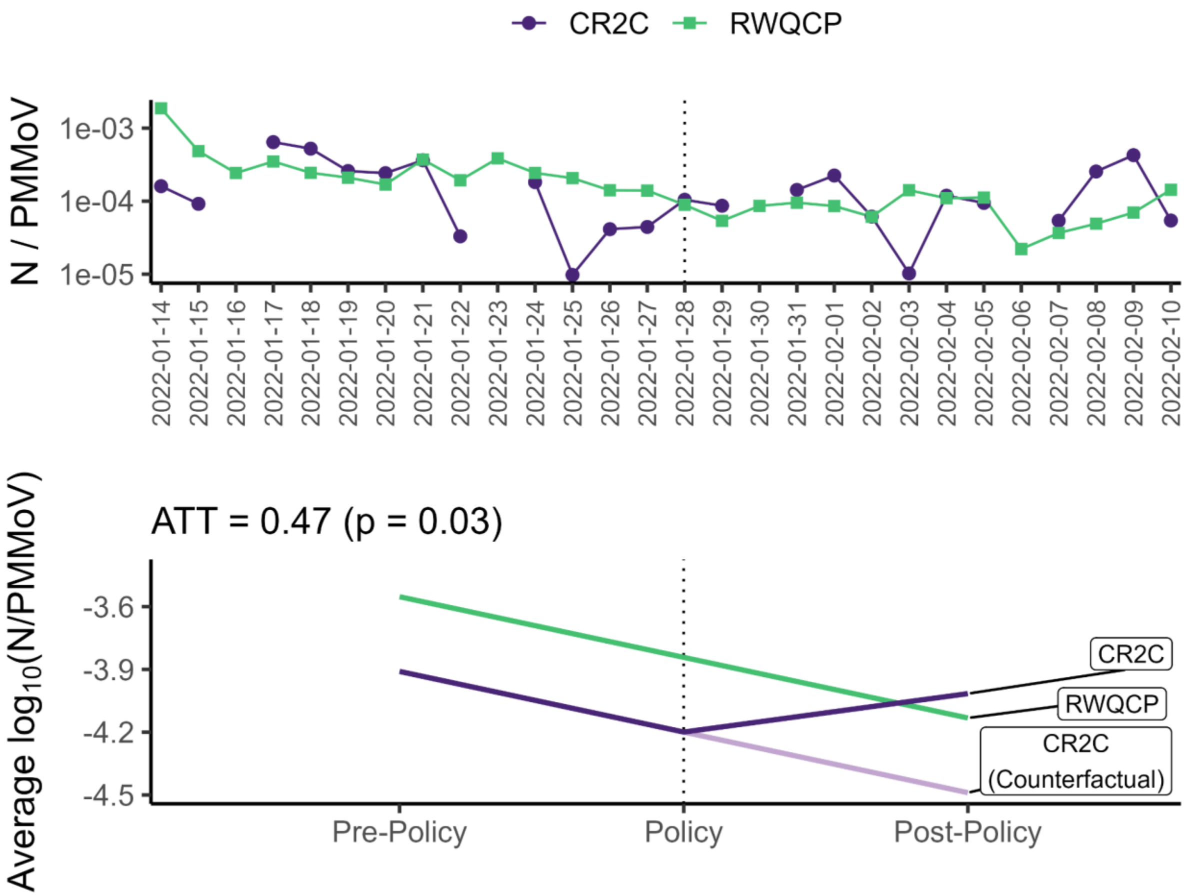


Figure S2. Depiction of the difference-in-differences (DiD) approach using the date when indoor events and gatherings were allowed to resume (28 January 2022) as an example. Top: Daily N/PMMoV concentrations at the Codiga Resource Recovery Center (CR2C) and Palo Alto Regional Water Quality Control Plant (RWQCP) over the 14 days before and after the policy change (denoted by the dotted line). Concentrations are displayed on a $\log_{10}$ scale. Bottom: Average $\log_{10}$(N/PMMoV) concentration at CR2C and RWQCP across the 14 days before and after the policy change. The counterfactual average $\log_{10}$(N/PMMoV) concentration at CR2C post-policy was estimated based on the time trend observed at RWQCP. The difference between the observed and counterfactual average $\log_{10}$(N/PMMoV) concentration at CR2C post-policy represents the average treatment effect on the treated (ATT). Here, a positive ATT value suggests that the policy change was associated with an increase in wastewater N/PMMoV concentrations at CR2C.

**Table S2. Difference-in-difference analysis for campus COVID-19 policies and events using two different comparison groups**

| Date (Trend/Level) | Policy or Event | Comparison: RWQCP<br><br>ATT (p value) | Comparison: RWF<br><br>ATT (p value) |
|---|---|---|---|
| 9/2/2021 (Upward/High) | Face coverings recommended outdoors in crowded settings; Indoor student parties prohibited | 0.72 (0.02) | 0.63 (0.03) |
| 9/20/2021 (None/Medium) | Surveillance testing required for faculty, staff, and postdoctoral scholars regardless of vaccination status; Revised travel guidelines in effect; Start of autumn quarter | -0.61 (0.05) | -0.62 (0.02) |
| 10/8/2021 (None/Low) | Indoor student parties can resume | -0.42 (0.2) | -0.42 (0.2) |
| 1/3/2022 (Upward/High) | Online classes start; Start of winter quarter | -0.078 (0.8) | 0.098 (0.8) |
| 1/5/2022 (Upward/High) | No indoor events and gatherings; Outdoor-only gatherings and meetings prohibited | -0.16 (0.6) | 0.045 (0.9) |
| 1/18/2022 (None/High) | In-person instruction resumes | -0.00048 (1.0) | -0.30 (0.2) |
| 1/21/2022 (None/High) | Outdoor-only gatherings and meetings allowed again | -0.11 (0.6) | -0.36 (0.1) |
| 1/28/2022 (Downward/Low) | Indoor events and gatherings can resume | 0.47 (0.03) | 0.34 (0.10) |
| 3/28/2022 (None/Medium) | Start of spring quarter | 0.47 (0.04) | 0.64 (0.001) |
| 4/7/2022 (None/High) | Surveillance testing no longer required for students | -0.62 (0.02) | -0.36 (0.1) |
| 6/12/2022 (None/Medium) | Commencement | 0.11 (0.6) | 0.15 (0.5) |
| 6/20/2022 (None/High) | Start of summer quarter | 0.34 (0.2) | 0.31 (0.2) |
| 9/26/2022 (None/Low) | Start of autumn quarter | 0.37 (0.09) | 0.37 (0.07) |
| 10/24/2022 (None/Low) | Masks no longer required in classrooms and on campus shuttles | 0.11 (0.7) | 0.072 (0.7) |
| 1/6/2023 | Face coverings strongly recommended indoors | -0.26 | -0.13 |

| | | | |
|---|---|---|---|
| (Downward/Medium) | and in crowded outdoor settings | (0.3) | (0.6) |
| 1/9/2023<br>(None/Low) | Start of winter quarter | 0.11<br>(0.6) | 0.023<br>(0.9) |
| 3/24/2023<br>(Upward/High) | Optional, free, laboratory-based PCR testing ends for employees | 0.72<br>(0.008) | 0.63<br>(0.01) |
| 4/3/2023<br>(None/High) | Start of spring quarter | -0.022<br>(0.9) | 0.036<br>(0.9) |
| 6/18/2023<br>(None/Low) | Optional, free, laboratory-based PCR testing ends for students; Commencement | 0.37<br>(0.3) | 0.40<br>(0.2) |
| 6/26/2023<br>(Upward/High) | Start of summer quarter | 0.32<br>(0.3) | 0.29<br>(0.3) |

RWQCP = Palo Alto Regional Water Quality Control Plant
RWF = San José-Santa Clara Regional Wastewater Facility
ATT = average treatment effect on the treated

We also implemented the difference-in-differences (DiD) analysis using wastewater data from the San José-Santa Clara Regional Wastewater Facility (RWF) as a comparison group (Table S2). RWF treats 110 million gallons of wastewater on average per day for several other communities throughout Santa Clara County.[12] Overall, we observed results that were similar to using the Palo Alto Regional Water Quality Control Plant (RWQCP) as a comparison group. All five policy change dates associated with a significant change (alpha ≤ 0.05) in wastewater N/PMMoV concentrations using RWQCP as a control were also associated with a significant change (alpha ≤ 0.05 or alpha ≤ 0.1) in wastewater N/PMMoV concentrations using RWF as a comparison group. There were no policy change dates associated with a significant change (alpha ≤ 0.05) in wastewater N/PMMoV concentrations using RWF as a control for which there was an insignificant change (alpha > 0.05) in N/PMMoV concentrations using RWQCP as a comparison. For the five policy change dates associated with a significant change in wastewater N/PMMoV concentrations, the sign of the average treatment effect on the treated (ATT) value was the same using either comparison sewersheds.
